# Endocrine, immune and disease dynamics in a patient with rheumatoid arthritis during flare and medication change

**DOI:** 10.1101/2024.03.18.24304149

**Authors:** Lennart Seizer, Johanna Gostner, Christoph Garbers, Melina Licht, Sebastian Sager, Christian Schubert

## Abstract

**Objective:** Rheumatoid arthritis (RA) is a chronic autoimmune disease of widely unknown etiology and pathophysiology. In this integrative single-case study on a patient with RA, we had the unique opportunity to closely monitor the individual dynamics of endocrine, immune and disease variables during a naturally occurring flare-up and subsequent medication change.

**Methods:** The female RA patient collected her entire urine over 30 days in 12-h intervals (60 consecutive measurements in total). Subsequently, interleukin-6 (IL-6), orosomucoid-2, cortisol (ELISA), neopterin and creatinine (HPLC) levels were determined in the urine samples. Further, each morning and evening, the patient completed the DIARI, a set of questionnaires on variables such as subjective pain, subjective RA disease activity and emotional states. Once a week, besides an online video interview, the patient had an appointment at her rheumatologist, in which several indices of RA disease activity were determined: SDAI, CDAI and DAS28. From these data various time series were constructed for statistical analysis.

**Results:** RA disease state increased from low to high activity during the first 12 study days. Thereupon, the medication was changed, which proved effective in reducing RA disease activity. However, the levels of urinary neopterin, urinary orosomucoid-2, and urinary IL-6 did not show any response, neither to the increasing disease activity nor the medication change. The patient’s daily reports on pain, RA disease activity, and emotional states, however, mirrored the course of the rheumatologic indices.

**Conclusion:** This integrative single-case study clearly demonstrated the importance of process analysis for the evaluation of therapeutic measures in RA. In the patient studied, urinary levels of neopterin, orosomucoid-2 and urinary IL-6 were not found to be appropriate biomarkers of short-term fluctuations in RA disease activity. Instead, the results reported by the patient proved to be a useful tool for ambulatory and longitudinal monitoring of RA.

## Introduction

Rheumatoid arthritis (RA) is a chronic autoimmune disease that includes synovial inflammation and hyperplasia, autoantibody production, cartilage, and bone destruction, and systemic features including cardiovascular, pulmonary, skeletal, and psychiatric disorders. The RA disease course is often characterized by fluctuations in disease activity with the sudden re-expression or enhancement of disease pathogenic processes, termed *flare*. For deeper insights into the concept of flares in RA see Bozzalla-Cassione et al. (2022). While biological processes during active RA are well known, there is little insight into the pathodynamics that accompany the transition from remission to flare-up, or whether remission in RA refers to the complete restoration of normal body functions (restitutio ad integrum) or to active regulatory mechanisms which keep pathologic processes in check (balanced homeostasis) (Bozzalla-Cassione et al., 2022). Further, despite significant progress in the treatment of RA, 20–30% of patients remain refractory to the state-of-the-art therapy (Smolen et al., 2020). Thus, key points in the European Alliance of Associations for Rheumatology’s (EULAR) research agenda contain the monitoring of therapeutic drug use and courses of disease to elucidate molecular pathways in treatment responses and identify associated biomarkers (Smolen et al., 2020).

This paper presents the results of an integrative single-case study designed to get insights into psychoneuroimmunological mechanisms of RA disease and therapy under the highest possible ecological conditions (“life as it is lived”) (Schubert et al., 2021). Specifically, over a period of one month, the patient collected her entire urine in pooled 12-h collections, completed questionnaires twice a day and had weekly online in-depth interviews. In addition, the patient had weekly appointments at her rheumatologist for RA activity assessment including blood sampling. Such N-of-1 designs have been recommended in clinical care research since they focus on individual, not average, responses to treatment and adequately handle within-subject variation (Schork, 2015; Seizer et al., 2023a). In the present study, it occurred by chance that the patient experienced an RA flare-up, which offered a unique possibility to investigate the emergence and termination of this flare with high temporal resolution of disease-related parameters under naturalistic conditions.

## Methods

### Study design

This integrative single-case study on a female RA patient is part of a larger trial on personalized therapy in RA (PETRA; Seizer et al., 2023b). Shortly before the start of the study, the patient was given a thorough physical examination and psychological assessment. Then, she collected her entire urine for 30 days in 12-h intervals from approx. 8 a.m. to 8 p.m. (day), and approx. 8 p.m. to 8 a.m. (night) using polyethylene urine containers (total of 60 12h-intervals). Upon collection, Na-Metabisulfite and Na-EDTA were added to the containers to prevent urine sedimentation and oxidation. At the end of each 12-h interval, the patient aliquoted the urine in 2,5 ml polypropylene tubes (Eppendorf) and froze the urine samples at -20°C. Once a week, the frozen samples were brought to the laboratory where they were stored at -70°C until further analysis (no breaking of the cold chain). Moreover, at the end of each 12-h interval (at approx. 8 a.m. and approx. 8 p.m.), the patient completed the Daily Inventory of Activity, Routine and Illness (DIARI), a set of questionnaires including two 100 mm visual analogue scales on subjective RA disease activity and pain as well as the short form of the *Eigenschaftswörterliste* (EWL), a German questionnaire that measures emotional states on three scales (mood, irritation, mental activity) (Becker et al., 1988). Furthermore, once a week, an in-depth interview was conducted per video with the patient to determine previous week’s emotionally meaningful positive and negative daily incidents. Also weekly, she had an appointment with her rheumatologist for blood sampling and to determine RA disease activity scores (total of 5 appointments). For further details on the study design, see Schubert et al. 2012.

### Subject description

The subject of this study was a Caucasian woman in her 50s. She was married, had one son and lived with her husband in southern Germany. She was diagnosed with Hashimoto’s thyroiditis about 15 years prior to study start, which was incidentally detected during a routine examination and was treated using a daily morning dose of thyroxine (37.5 mg). About 5 years prior to study start, she was diagnosed with RA. Additionally, during the last three years, the patient complained about having headaches and problems sleeping through. At study entry her medication plan regarding RA consisted of a daily morning dose of prednisolone (1-2 mg) and sulfasalazine (2 g).

### Urine measurements

Urinary creatinine and neopterin concentrations were measured using High-Pressure Liquid Chromatography (HPLC) (Model LC 550; Varian Associates, Palo Alto, CA). Urinary cortisol (NT-DNOV010, Biomedica), ORM-2 (ABIN6969086) and IL-6 (DY206, R&D Systems) were determined using enzyme-linked immunosorbent assays (ELISA). Cortisol is the main effector of the hypothalamic-pituitary-adrenal (HPA) axis and has well described immunomodulatory properties (Cain and Cidlowski, 2017). Neopterin is produced by monocytes and macrophages upon stimulation of T helper type 1 (th1)-cell derived gamma-interferon (IFN-γ) and is used as a marker of non-specific systemic inflammation. Urinary levels represent neopterin biosynthesis well as it is not biologically active, does not bind to receptors and undergoes rapid renal clearance (Stuart et al., 2020). IL-6 is a pleiotropic cytokine with diverse functions in coordinating innate and adaptive immune responses (Srirangan & Choy, 2010). So far, there is no definitive answer on the proportionality of urine and serum IL-6 levels. Although the kidneys take part in the (urinary) elimination of IL-6 from systemic circulation with a pass ratio of 0.2% (Nowak et al., 2012), urine and serum levels did not correlate in a cross-sectional analysis of spot samples (Nobles et al., 2015). ORM-2 is an acute-phase reactant and its urinary levels have previously been identified as surrogates for assessing disease activity and prognosis of RA (Park et al., 2016). For determination of each of these biomarkers, all 60 consecutive urine concentrations were measured in one single run, at least two times independently, and each time using a new aliquot. Afterwards, the results for each of the biomarkers were averaged. Urinary neopterin, IL-6, ORM-2 and cortisol levels are expressed in relation to urinary creatinine (per mol) to compensate for variations in urine density. As the use of creatinine to normalize urinary biomarkers has been questioned previously, we additionally considered 12-h urine excretion rates (amount of substance in test volume * total 12-h urine volume) for normalization (Waikar et al., 2010).

### Rheumatic disease activity

Rheumatic disease activity over the study period was assessed using a combination of standard indices and patient-reported measures. Twice a day (at approx. 8 a.m. and approx. 8 p.m.), the patient completed subjective appraisals on RA disease activity and pain using 100 mm visual analog scales (VAS) (Fautrel et al., 2018). Moreover, once a week, the patient had an appointment with her rheumatologist in which the clinical disease activity index (CDAI) and the simplified disease activity index (SDAI) were determined. In addition, levels of C-reactive protein (CRP) in plasma and erythrocyte sedimentation rate (ESR) were analyzed to determine the disease activity index-28 score CRP (DAS28-CRP) and ESR (DAS28-ESR) (Aletaha & Smolen, 2005; Bray et al., 2016). The patient met her rheumatologist at 12-h intervals 1, 10, 24, 38 and 52. Cut-off values for SDAI, CDAI and DAS28 to determine different RA disease activity states were used as proposed by Aletaha & Smolen, 2005.

### Statistical analysis

All statistical analyses were performed using *R* 4.2. Pearson product-moment correlations were conducted to determine the similarity between different normalization procedures of the urinary biomarkers. To investigate serial dependencies in the time series, autocorrelation functions (ACF) were calculated. Two-sided, non-paired t tests were performed to evaluate the difference in time series before and after the medication change. In all analyses, statistical significance was considered at *p* < .05.

## Results

The patient experienced an increase in RA disease activity during the study period as determined by SDAI, CDAI, and DAS28-CRP/-ESR. Thereupon, her medication plan was adjusted (at 12-h interval number 25): the daily morning dose of prednisolone was elevated from 1-2 mg to 5 mg, sulfasalazine was deposed, and a weekly injection of methotrexate (7.5 mg) was introduced (methotrexate injections were given at 12-h intervals 24, 38, and 52). All the weekly measures of RA disease activity that were assessed by the rheumatologist (CDAI, SDAI, CRP, ESR, DAS28-CRP/-ESR) showed a similar (inversed U-shaped) temporal pattern: an increase during the first 12 days of the study (until 12-h interval number 24) and a decline after the change in medication (Fig. 1).

**Figure 1:**
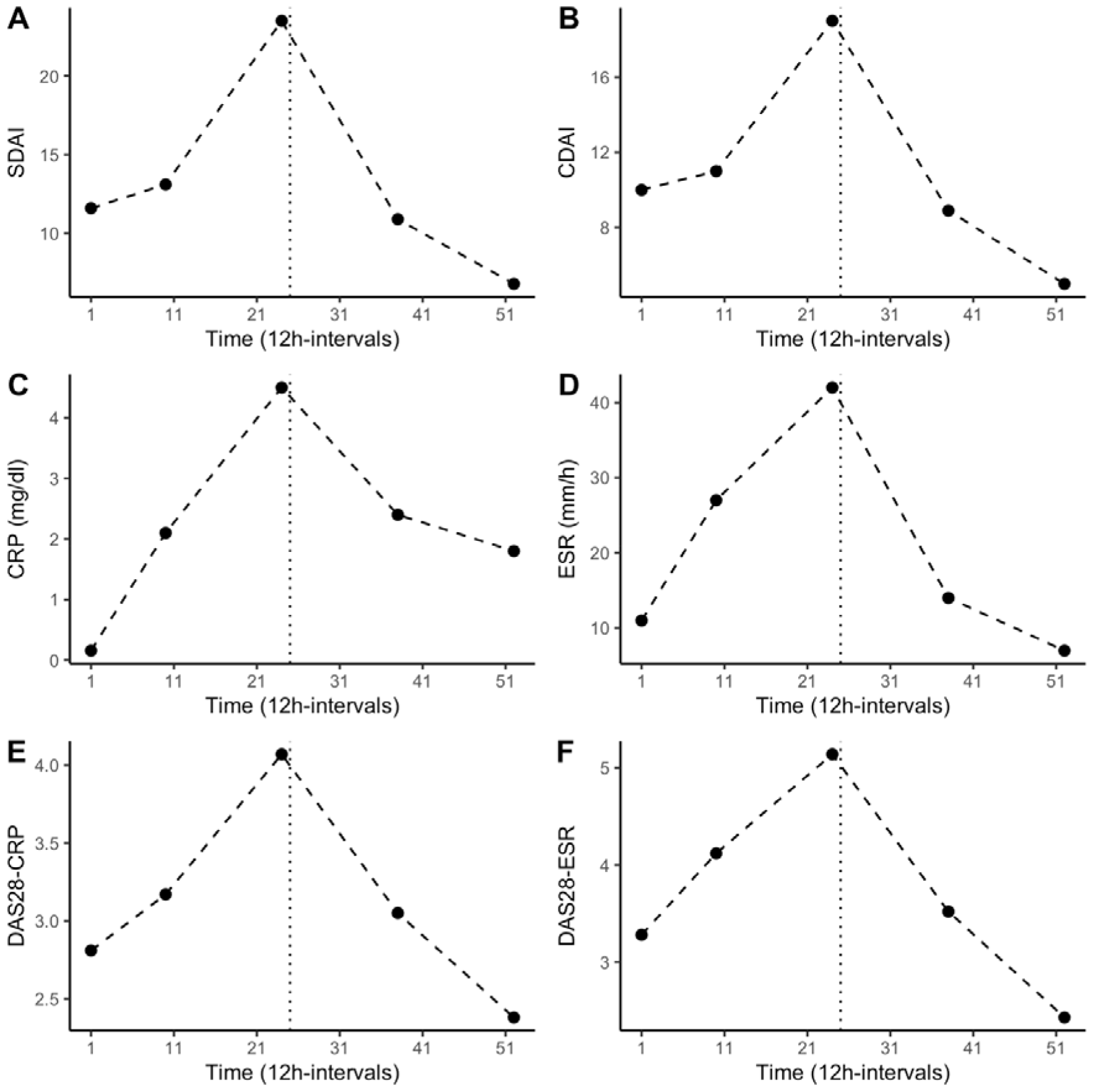
Weekly determinations of the patient’s RA disease activity by the rheumatologist at 12h-intervals 1, 10, 24, 38 and 52. The dotted vertical line marks the timepoint of change in medication. (A) Simplified disease activity index (SDAI), (B) clinical disease activity index (CDAI), (C) c-reactive protein (CRP), (D) erythrocyte sedimentation rate (ESR), (E) disease activity score-28 (DAS28) using CRP, (F) DAS28 using ESR.

Descriptive statistics on the consecutive urinary 12-h measurements (creatinine, cortisol, neopterin, ORM-2, IL-6, and the patient-reported outcomes) are listed in Table 1, while their time-series are plotted in Fig. 2. The resulting time series from the different normalization procedures were significantly correlated for each biomarker. For completeness, all the following statistical analyses were repeated using time series with 12-h urine excretion rates as alternatives to creatinine-normalized values, but no difference in the directions or significances of the results emerged (data not shown).

**Table 1:**
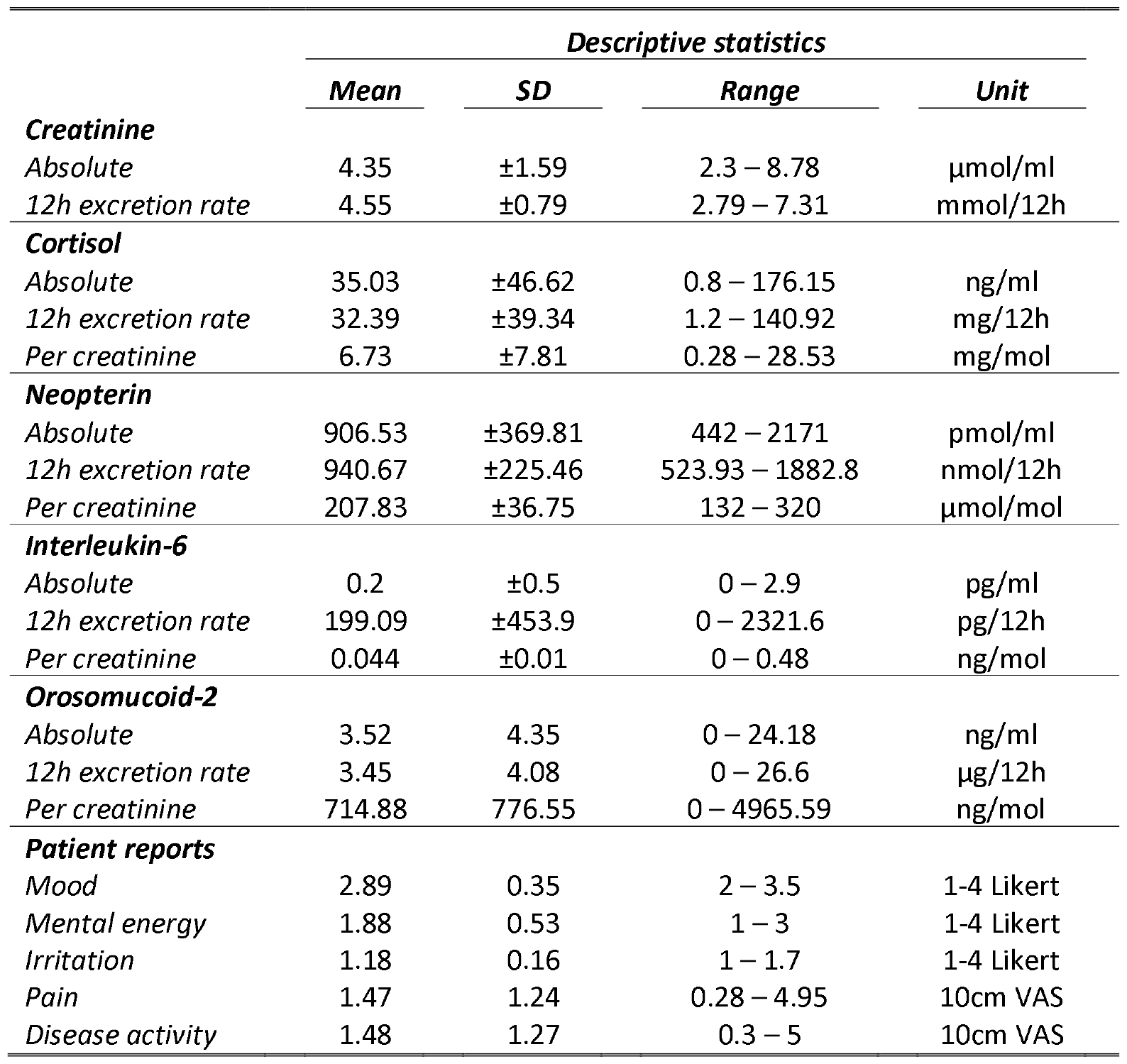
Descriptive statistics for the levels of the 12h-urine analytes in their absolute concentration (not normalized), in their 12h excretion rate (concentration * total 12h urine volume) and per creatinine (analyte concentration / creatinine concentration).

**Figure 2:**
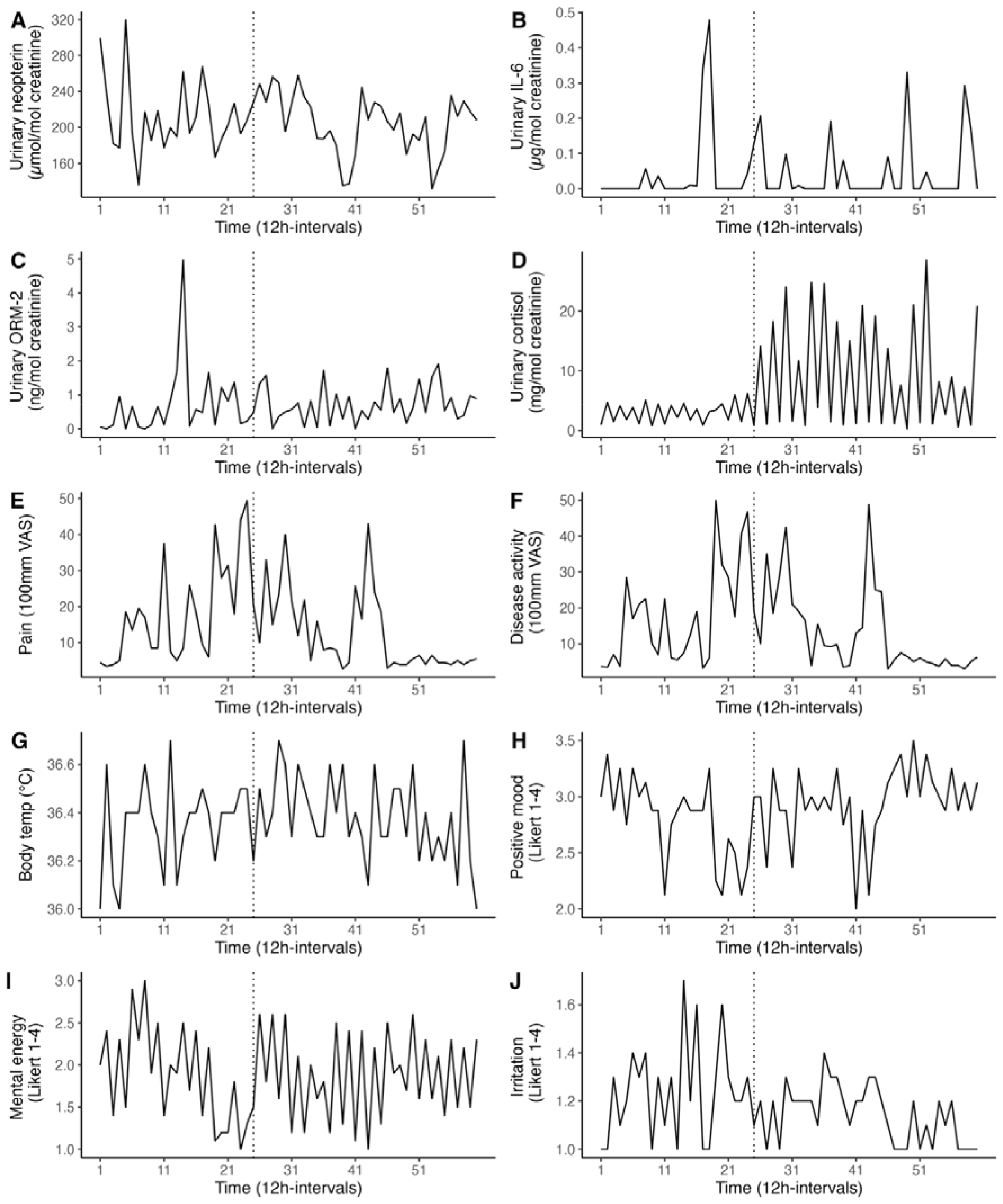
Various time series covering the 30-day study period in 12h-intervals (T = 60). The dotted vertical line marks the timepoint of change in medication. (A) Urinary neopterin, (B) urinary interleukin-6 (IL-6), (C) urinary orosomucoid-2 (ORM-2), (D) urinary cortisol, (E) subjective pain, (F) subjective RA disease activity, (G) body temperature, (H) mood, (I) mental activity, (J) irritation.

A significant negative ACF at Lag1 was found in urinary cortisol levels (*r* = -.47, *p* < 0.01), indicating a circadian pattern with higher values during the day (12.05 ± 8.05 mg/mol creatinine) and lower values during the night (1.4 ± 0.77 mg/mol creatinine). Urinary IL-6, urinary ORM-2 and urinary neopterin concentrations did not exhibit significant circadian rhythms.

To evaluate differences in the urinary 12-h measurements before (12-h intervals 1-24) and after (12-h intervals 25-60) the change in medication, two-sided, non-paired t-tests were performed. Urinary cortisol levels increased significantly after the change (*t* = 3.91, *p* < 0.01). However, this increase was only evident for day levels (*t* = 3.88, *p* < 0.01), but not for night levels (*t* = 0.38, *p* = 0.71). The levels of neopterin (*t* = 0.64, *p* = .52), IL-6 (*t* = -0.19, *p* = .85), ORM-2 (*t* = -0.09, *p* = .93), subjective pain (*t* = 1.67, *p* = .10), subjective RA disease activity (*t* = 1.34, *p* = .19), mood (*t* = -1.68, *p* = .10), irritation (*t* = 2.09, *p* = .05), mental activity (*t* = 0.09, *p* = .93), and body temperature (*t* = -.40, *p* = .63) did not differ before and after the medication change. A polynomial regression analysis was conducted to replicate the non-linear pattern in the disease activity indices (Fig. 1). Thereby, a linear and quadratic term of time was fitted to the time series. Subjective pain, subjective disease activity, mood, irritation, and body temperature showed a U-shaped pattern, mirroring the disease activity indices (Fig. 2). No such pattern was found for the time-series of urinary IL-6, urinary ORM-2, urinary neopterin and urinary cortisol levels.

## Discussion

In this study, we monitored endocrine, immune and disease activity parameters in an RA patient over 30 days in 12h-intervals during a flare-up and subsequent change in medication.^1^ During the study period, the disease states increased from low to high activity during the first 12 days (as measured by SDAI, CDAI and DAS28-CRP/-ESR). Thereupon, the medication plan was changed: the daily morning dose of prednisolone was elevated from 1-2 mg to 5 mg, sulfasalazine was deposed, and a weekly injection of methotrexate (7.5 mg) was introduced. This change in medication proved successful in reducing the RA disease activity towards scores considered to indicate low activity or remission within two weeks (Aletaha & Smolen, 2005) (Fig. 1). Previously, CRP and ESR have been found to differ in their usefulness to monitor RA disease activity and treatment response, as CRP levels tend to drop quickly with treatment, while ESR levels can take weeks to normalize (Bray et al., 2016). Here however, when applying naturalistic time series data, no difference was found in the responsive delays of CRP and ESR (Fig. 1 C and D).

Urinary cortisol levels increased significantly with the new medication regime but likely did not represent enhanced endogenous cortisol production but rather originated as a product of the ELISA-kit’s cross reactivity with prednisolone (46%; Fig. 2 D). However, only the diurnal levels of cortisol increased, whereas the nocturnal levels did not differ pre-post medication change, suggesting that the basal circadian rhythm was unaffected by this medication. The urinary assessments of immune system activity (IL-6, ORM-2, neopterin) were neither associated to the flare-up nor the change in medication, but instead remained stationary throughout the study period (Fig. 2 A, B and C). Only two decreases (12h-intervals 39 and 53) can be seen in the time series of neopterin after the change, which are likely explained by the newly introduced weekly methotrexate injections administered directly before these neopterin decreases during 12h-intervals 38 and 52.

These data indicate that urinary neopterin measurements are unrelated to short-term changes in disease activity and acute exacerbations. Such finding is consistent with a recent meta-analysis examining the role of neopterin in RA, which concludes that while neopterin levels are generally elevated in RA patients, no relation exists between specific neopterin levels and states of disease activity (Hejrati et al., 2020). This points to a difference in the biological pathways of RA-associated inflammation as measured by CRP and ESR, and neopterin-associated systemic inflammation. Since IL-6 and ORM-2 levels also did not correspond to the flare-up in our study, the same considerations might apply for these biomarkers as well. However, as IL-6 has previously been described as a crucial part in the immunological pathways of RA (Srirangan & Choy, 2010), the non-existent link between IL-6 and RA disease activity might also result from a limited comparability between serum and urine levels (Nobles et al., 2015; Nowak et al., 2012).

This study’s limitations emerge primarily from the fact that the described results were generated in only one patient with RA limiting generalizability to the population. Moreover, assessment of neopterin, ORM-2 and IL-6 in the blood or synovial fluid of the patient additionally to the urinary measurements could have given more insights into their role and value as biomarkers. Also, the use of advanced time-series analysis, such as vector autoregression models, could give further insights into the temporal relations among variables in future studies.

In conclusion, this integrative single-case study clearly demonstrated the importance of process analysis for the evaluation of therapeutic measures in RA. In the patient studied, urinary neopterin, urinary ORM-2 and urinary IL-6 did not represent valid biomarkers for short-term changes in disease activity of RA. On the other hand, the results also indicate, in accordance with previous reports (Bozzalla-Cassione et al., 2022; Fautrel et al., 2018), that patient-reported outcomes and emotional states can be a useful tool in the ambulatory and longitudinal monitoring of disease activity, flare occurrence and treatment responses. Overall, this N-of-1-study highlights the benefit of frequently collecting psychological and immunological data over an extended period of time under conditions, which preserve the natural ebb and flow of daily life, with no external variability created by study-related guidelines or restrictions.

## Data Availability

All data produced in the present study are available upon reasonable request to the authors

## Acknowledgements

We would like to thank the patient of this study for her participation.

## Footnotes

1 The urine samples generated in this study are quite unique, as they allow the flare-up of RA disease activity and subsequent therapeutic response to be tracked continuously in a setting of high ecological validity. Multiple aliquots of each 12-interval’s urine collection are stored at –80°C. We would like to share this biobank with other researchers for further analysis and hypothesis-testing of the role of biomarkers in short-term changes of disease activity in RA.

